# Evaluation of The Everyday Memory Questionnaire-Revised in Menopausal Population: Understanding the Brain Fog During Menopause

**DOI:** 10.1101/2023.03.14.23287272

**Authors:** Chen Zhu, Elizabeth HX Thomas, Qi Li, Shalini Arunogiri, Natalie Thomas, Caroline Gurvich

**Author notes:** Address correspondence to: Caroline Gurvich, HER Centre Australia, Department of Psychiatry, Central Clinical School, Faculty of Medicine, Nursing and Health Sciences, Monash University and The Alfred Hospital, Melbourne, Australia.

## Abstract

**Introduction:** Brain fog (i.e., memory complaints and concentration difficulties) is frequently reported during the menopausal transition. There is lack of standardized scales available to measure brain fog across the menopause transition. This study aimed to evaluate the factor structure of the Everyday Memory Questionnaire-Revised (EMQ-R) and to determine the most commonly reported everyday cognitive symptoms in a menopausal population.

**Methods:** 417 females, including 107 pre-menopausal, 149 peri-menopausal, and 161 early post-menopausal, met criteria and were included in the analyses. Memory and attention related symptoms were measured using the EMQ-R. Confirmatory factor analysis (CFA) was conducted to test the model fit of the bifactor structure (attentional and retrieval) of the EMQ-R in menopausal populations and evaluated by Cronbach’s alpha test. One way ANOVA and MANCOVA were used to investigate the group differences of individual items and two subscales.

**Results:** CFA indicated that the bifactor model (*retrieval* and *attentional*) of the EMQ-R has a good fit in all three groups. A significant difference (with the highest mean score observed in peri-menopausal group) was identified in the retrieval subscale score (*F*=3.17, *p*=0.043) but not in the attentional subscale or total scores amongst the three groups. Items (1, 3, 5) related to daily information retrieval problems were significantly reported in the peri-menopausal group.

**Discussion:** The EMQ-R serves as an instrument to measure memory and attention symptoms, referred to as ‘brain fog’ in menopause. Increased memory retrieval complaints reported by peri-menopausal group suggests a transition-related memory retrieval dysfunction during menopausal transition.

## Introduction

Everyday memory complaints and difficulty concentrating are frequently reported during the menopause transition. Up to 70% of women report attention and memory problems during menopause combined with other menopausal symptoms, such as hot flashes, depressed mood, and night-time awakening.^1-4^ Poor memory and poor concentration during menopause can have a significant impact on work and life in general.^5^ In addition, associations between subjective complaints and objective measurements of cognition further support the women’s accurate appraisals of their cognitive complaints experienced in daily life and their influences on daily functioning.^3,6-8^ Other than objective measures of cognition^6,9^, subjective cognitive problems are also predicted by lifestyle factors (e.g., dietary habit, physical exercise), psychosocial variables (e.g., anxiety, depression, attitude towards menopause), and vasomotor symptoms (e.g., hot flashes, night sweats, sleep disturbance).^2,3,10^

The term ‘brain fog’ has been widely used in recent years in both popular literature and scientific research to describe subjective cognitive symptoms during the menopause transition. Consistent with the recent work^11,12^, this paper defines the brain fog as the constellation of subjective cognitive symptoms reported by women with a particular focus on memory and attention complaints. Currently, there is no standardized measurement of menopausal-related brain fog and only one available study adopted the term ‘brain fog’ as the research outcome which was roughly assessed by the Mini-Mental State Exam with a higher score indicating lower likelihood of brain fog.^12^ Instead of examining the brain fog related memory and attention problems independently via various, often unstandardized, questionnaires, identifying a validated and standardized questionnaire to assess brain fog in menopausal population is necessary.

### Subjective Measurement of Cognition

Several questionnaires have been employed to measure subjective experiences of cognitive change during menopause, The Memory Functioning Questionnaire (MFQ) has been in USA studies^6,13^, and the Multifactorial Memory Questionnaire (MMQ) was used in an Australian sample^14^. Both of these questionnaires were initially developed for healthy older adults^15,16^ and were later validated and evaluated in other populations, such as epilepsy^17^, Multiple Sclerosis^18^ and Dutch older adults^19^, but not specifically validated in a menopause population. In addition to validated questionnaires, some menopause studies used a single ‘yes’ or ‘no’ question (e.g., ‘Do you have any memory problems compared to the past’ or ‘Do you have problems with attention/concentration’) to measure subjective memory or attention problems.^3,20,21^ Given the lack of validation studies and diverse measurements used to measure subjective cognition in previous menopausal samples, it is important to identify a standardized measurement of subjective cognition (i.e., brain fog) targeting both memory and attention problems to allow a more direct comparison and justified summary of findings across different studies.

The Everyday Memory Questionnaire-Revised (EMQ-R) is a subjective measure of memory failure in everyday life, which was initially developed as a 35-item questionnaire to investigate the effects of closed-head injuries on memory performance.^22^ Subsequently, this questionnaire was reduced by Royle and Lincoln ^23^ to a 13-item one (EMQ-R) and validated in several clinical populations, such as stroke^24^, chronic pain^25^ and HIV^26^ and non-clinical populations, such as community younger and older adults^27^, and Bosnian/Serbian healthy adults^28^. Although the item loadings were slightly different in these studies, two main factors of the EMQ-R were consistently observed with one factor relating to memory retrieval and another focusing on attentional process, suggesting that the EMQ-R may serve as a better option to measure brain fog due to this bifactor structure (*Retrieval* and *Attentional*).

Comparing with other instruments, such as the MFQ consisting of 64 items and a few outdated descriptive statements (e.g., keep an appointment book), the EMQ-R was validated more recently with a relatively short questionnaire that can be easily implemented in survey or everyday life and decrease the drop rate of the questionnaire. Moreover, unlike the four memory-related factors (i.e., general frequency of forgetting, seriousness of forgetting, retrospective functioning, and mnemonics usage) identified in the MFQ^15^, the EMQ-R may constitute a more appropriate measurement of subjective cognition in menopausal population given its unique *attentional* factor that can potentially help understand the interactions between subjective memory and attention.

Thus, the goal of this study was to evaluate the factor structure (internal consistency) of the Everyday Memory Questionnaire-Revised (EMQ-R) in menopausal population. Based on Royle and Lincoln ^23^, a bifactor structure (*Retrieval* and *Attentional*) was expected in the current study. Moreover, the current study also aimed to look at the difference in subjective cognition across different menopausal stage as well as to determine the prevalence and most commonly reported everyday cognitive symptoms in menopausal women based on items and subscales from the EMQ-R, so that to further help women and clinicians better understand the brain fog in menopause.

## METHODS

### Participants

This research was approved by the Monash University Human Research Ethics Committees (Ref. 2021-27653-66980). Online and social media advertisements, from November 2021 to August 2022, promoted the research study and sought to identify women who were potentially perimenopausal and potentially experiencing symptoms, such as brain fog, hot flashes, and night sweats. Interested women were then preliminarily screened based on four initial criterions: a) Aged between 35 and 60 years (to better capture the early/late menopausal population); b) Being biologically female; c) Fluent in English; and d) Not pregnant or lactating. Exclusion criteria included severe health conditions or neuropsychiatric disorders that significantly impact cognition, unable to give informed consent, or unable to demonstrate understandings of the objectives of the study and the questionnaires.

### Procedure

The data was collected as part of a larger Menopause and Cognition online study (Meno-COG), which was managed using REDCap hosted at Monash University.^29,30^ REDCap is a secure, web-based and confidential platform for the collection of research data allowing flexible data manipulations and export procedures. Preliminary screening was firstly delivered online via REDCap. Once eligibility was confirmed, explanatory statement and consent form were sent to participants for reading and providing electronic signature. Participants were then required to complete self-reported questionnaires regarding their demographic, socio-economic, lifestyle and medical conditions. Menopausal related information was also collected including the current and previous use of hormonal treatment, the characteristics of menstrual cycle, number of parities, and views about menopause. Questions related to menstrual cycle pattern (e.g., Do you currently experience regular menstrual cycles? When did your cycles become irregular? When was your last menstrual period?) were generated based on the Stages of Reproductive Aging Workshop (STRAW) ^31^ to help define the menopausal stag. After completing questionnaires of basic information, participants were then directed to complete the EMQ-R.

### Outcome measure

The EMQ-R was used to measure brain fog in this study.^23^ It was a 13-item questionnaire where each item is a statement describing daily memory (e.g., Having to check whether you have done something that you should have done) or attention (e.g., When reading a newspaper or magazine, being unable to follow the thread of a story; losing track of what it is about) related behaviors (see Supplementary Table for detailed information). Participants were required to rate how often on average they think the item happened over the past month on a 5-point scale, with 0 = once or less in the last month, 1 = more than once a month but less than once a week, 2 = about once a week, 3 = more than once a week or less than once a day, 4 = once or more in a day. The total EMQ score (ranging from 0 to 52), and two subscale scores, retrieval (ranging from 0 to 28) and attentional (ranging from 0 to 15), were calculated by summing up the corresponding items, with a higher score indicating greater presence of brain fog, memory, or attention difficulties.

### Data Analysis

The detailed screening process was shown in Figure 1. Based on the characteristics of menstrual cycle (e.g., time of the last menstrual period, the regularity of menstrual cycles), participants were stratified into pre-menopausal, peri-menopausal and early post-menopausal groups according to the STRAW. One-way ANOVA and non-parametric tests (Kruskal-Wallis rank sum test) were used to explore the significant differences in demographic variables amongst the three groups, if applicable.

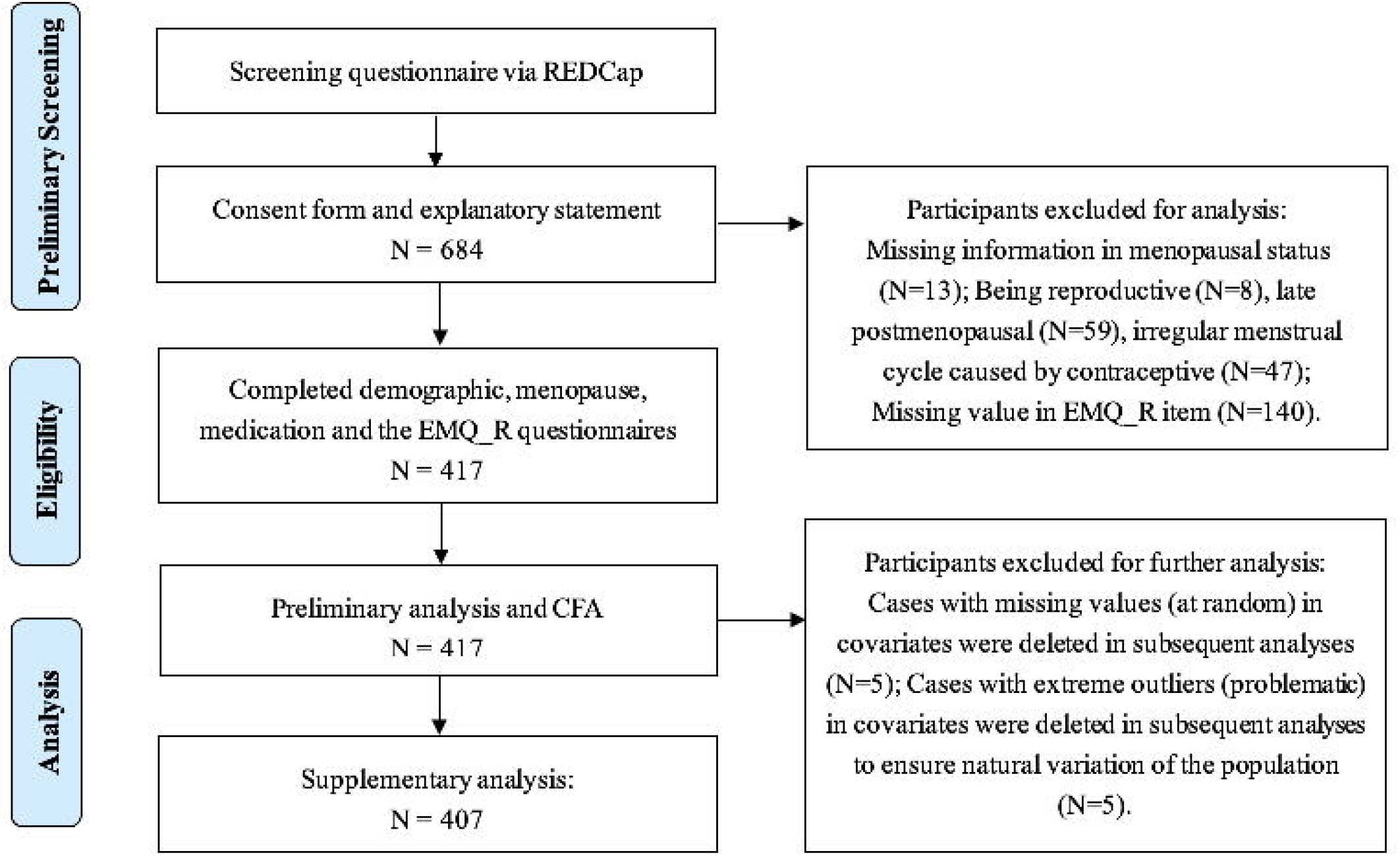

Confirmatory Factor Analysis (CFA) is often used to test how well a hypothesized measurement model fits a sample from a different population.^32^ The hypothesized EMQ-R factor model in this study was referred to the bifactor model identified in Royle and Lincoln^23^, which were the factor 1-*attentional* (items 8,9,11,13) and factor 2 – *retrieval* (items 1,2,3,5,6,7,10). Item 4 and item 12 were not included and classified into any individual factors. Lavaan, a package in R (https://lavaan.ugent.be), was employed to evaluate the fit of proposed EMQ-R factor model in pre-, peri- and early post-menopausal populations separately by Chi-square statistic (*X*^2^). Model reliability and strength were further explored and validated through Cronbach’s alpha test (α).

The relationships between the EMQ-R related scores and demographic data were investigated by either Pearson Correlation Coefficient or ANOVAs, where demographic variables that significantly correlated with the two subscale scores were used as a covariate. One way ANOVA and MANCOVA were used to investigate the difference in individual EMQ-R items as well as two subscales across three groups and determine the most commonly reported everyday cognitive symptoms in menopausal population. The p values were corrected through Bonferroni in multiple comparisons between each group to better understand the effect trend. In this study, statistical significance was set at the level of p < 0.05. All the analyses were performed in IBM SPSS Statistics (Version 25) and R (Version 4.2.1, https://www.r-project.org/).

## RESULTS

In total, 684 women initiated the survey, of which 417 women (107 Pre-menopausal, 149 Peri-menopausal, and 161 Early post-menopausal) aged 41 to 61 years had complete data and were included in the confirmatory factor analysis and group analysis, and 407 women were included in the multi-group comparison.

### Sample characteristics

Descriptive information of demographic variables and differences among groups were summarized in Table 1. Age was significantly different across the three groups, indicating an increasing trend with the progression of menopausal stage. All other variables were not significantly different across the groups. Descriptive analysis results of medical history were displayed in Table 2, without any significant difference identified amongst the three groups.

**TABLE 1.** Descriptives of Demographic Information

| Variable | Pre (N=107) | Peri (N=149) | Early-post (N=161) | p-value | Test |
| --- | --- | --- | --- | --- | --- |
|  | Mean (SD) | Mean (SD) | Mean (SD) |  |  |
| Age | 49.42 (2.72) | 51.17 (3.56) | 53.48 (3.83) | < **0.001\*** | ANOVA |
| No. of Parity | 2.46 (1.64) | 2.34 (1.59) | 2.25 (1.59) | 0.563 | ANOVA |
| BMI_Score | 27.17 (7.14) | 26.91 (6.82) | 26.64 (6.24) | 0.814 | ANOVA |
| *BMI_Category* | N = 106 | N = 145 | N = 161 | 0.094 | *Kruskal-Wallis rank sum tests* |
| Underweight (BMI<18.5) | 0 | 1 | 7 |  |  |
| Normal (18.5≤BMI<25) | 46 | 71 | 68 |  |  |
| Overweight (25≤BMI<30) | 37 | 37 | 50 |  |  |
| Obese (BMI≥30) | 23 | 36 | 36 |  |  |
| *Marital Status* | N = 105 | N = 148 | N = 161 | 0.396 | *Kruskal-Wallis rank sum tests* |
| Never Married | 16 | 22 | 15 |  |  |
| Married/Civil Partnership | 67 | 105 | 119 |  |  |
| Divorced/Seperated | 21 | 20 | 25 |  |  |
| Widowed | 1 | 1 | 2 |  |  |
| *Ethnicity_Category* | N = 106 | N = 148 | N = 161 | 0.296 | *Kruskal-Wallis rank sum tests* |
| African | 2 | 1 | 1 |  |  |
| Americas | 4 | 8 | 1 |  |  |
| Americas/African | 0 | 0 | 1 |  |  |
| Asian | 6 | 6 | 1 |  |  |
| European | 37 | 62 | 72 |  |  |
| European/African | 1 | 0 | 0 |  |  |
| European/Americas | 1 | 1 | 0 |  |  |
| European/Asian | 0 | 0 | 1 |  |  |
| Oceanian | 50 | 68 | 80 |  |  |
| Oceanian/European | 5 | 2 | 4 |  |  |
| *Education_Category* | N = 107 | N = 148 | N = 161 | 0.640 | *Kruskal-Wallis rank sum tests* |
| Secondary School (incomplete) | 0 | 2 | 2 |  |  |
| Secondary School (complete) | 12 | 7 | 15 |  |  |
| Tafe or Trade | 15 | 22 | 23 |  |  |
| Bachelor degrees | 41 | 62 | 70 |  |  |
| Masters | 32 | 45 | 35 |  |  |
| Doctorate | 7 | 10 | 16 |  |  |
| *Caffeine_Daily* | N = 107 | N = 149 | N = 161 | 0.513 | *Kruskal-Wallis rank sum tests* |
| Heavy drinkers (3 or more servings) | 41 | 74 | 82 |  |  |
| Mild_moderate drinkers (1-2 servings) | 61 | 58 | 58 |  |  |
| Non_drinkers | 5 | 17 | 21 |  |  |
| *Alcohol Intake* | N = 107 | N = 149 | N = 161 | 0.578 | *Kruskal-Wallis rank sum tests* |
| Never | 13 | 22 | 20 |  |  |
| Monthly | 28 | 32 | 38 |  |  |
| 2-4 times a month | 19 | 32 | 26 |  |  |
| 2-3 times a week | 36 | 47 | 51 |  |  |
| 4 or more times a week | 11 | 16 | 26 |  |  |

N = number of participants, SD = Standard Deviation.
\* indicated $p < 0.05$ .

**TABLE 2.** Descriptives of Medical Information

| Variable | Pre<br>(N=107) | Peri<br>(N=149) | Early-post<br>(N=161) | *p*-value | Test |
| --- | --- | --- | --- | --- | --- |
| *Current Psychiatric Therapy* | N=106 | N=149 | N=160 | 0.234 | *Kruskal-Wallis rank sum tests* |
| No | 85 | 131 | 132 |  |  |
| Yes | 21 | 18 | 28 |  |  |
| *Current Psychiatric Medication* | N=107 | N=147 | N=159 | 0.910 | *Kruskal-Wallis rank sum tests* |
| ADHD | 0 | 1 | 1 |  |  |
| Antidepressant (Other) | 2 | 3 | 3 |  |  |
| Antidepressant (SNRIs) | 5 | 4 | 9 |  |  |
| Antidepressant (SNRIs)&ADHD | 0 | 1 | 0 |  |  |
| Antidepressant (SSRIs) | 15 | 16 | 14 |  |  |
| Antidepressant (SSRIs&Other) | 1 | 0 | 1 |  |  |
| Antidepressant (SSRIs)&Benzodiazepine | 0 | 1 | 0 |  |  |
| Antidepressant (SSRIs)&Other | 0 | 0 | 1 |  |  |
| Not taking | 84 | 121 | 130 |  |  |
| *Current Hormonal Therapy* | N=106 | N=147 | N=155 | 0.234 | *Kruskal-Wallis rank sum tests* |
| Hormonal Contraceptives (HC) | 9 | 6 | 3 |  |  |
| Menopausal Hormone Therapy (MHT) | 1 | 33 | 53 |  |  |
| HC&MHT | 26 | 0 | 1 |  |  |
| Not taking | 67 | 107 | 93 |  |  |
| Other | 3 | 1 | 5 |  |  |
|  | 1 | 2 | 6 |  |  |
| *Diagnostic Psychiatric Disorder* | N=106 | N=149 | N=161 | 0.871 | *Kruskal-Wallis rank sum tests* |
| ADHD&ASD | 0 | 0 | 1 |  |  |
| ADHD&Mood disorder | 1 | 1 | 1 |  |  |
| ADHD&PTSD&Mood disorder | 0 | 0 | 1 |  |  |
| ASD&Mood disorder | 0 | 0 | 1 |  |  |
| Mood disorder | 23 | 35 | 35 |  |  |
| Mood disorder&Eating disorder | 0 | 0 | 1 |  |  |
| Mood disorder&PTSD | 0 | 0 | 1 |  |  |
| Non diagnosis | 80 | 112 | 119 |  |  |
| PMDD | 1 | 1 | 0 |  |  |
| PMDD&Mood disorder | 0 | 0 | 1 |  |  |
| PTSD | 1 | 0 | 0 |  |  |

*N* = number of participants.
ADHD, Attention-deficit/hyperactivity disorder; SNRIs, Serotonin and norepinephrine reuptake inhibitors; SSRIs, Selective serotonin reuptake inhibitors; ASD, Autism spectrum disorder; PTSD, Post-traumatic stress disorder; PMDD, Premenstrual dysphoric disorder.

### Confirmatory factor analysis

The hypothesized bifactor model (*Retrieval*: including item 1,2,3,5,6,7,10 and *Attentional*: including item 8,9,11,13) was represented in Figure 2. The significant result suggested the good fit of proposed models in all three groups: Pre *X*^2^(63, 107) = 146.05, *p* < 0.001; Peri *X*^2^(63, 149) = 156.26, p < 0.001; Early post *X*^2^(63, 161) = 194.67, *p* < 0.001. The Cronbach α for each subscale with 95 percent confidence interval were shown as follows, suggesting good internal consistency: *Retrieval*: Pre α = 0.88 [0.832 0.908] Peri α = 0.86 [0.818 0.887], Early post α = 0.88 [0.844 0.908]; *Attentional*: Pre α = 0.77 [0.653 0.841], Peri α = 0.77 [0.692 0.830], Early post α = 0.79 [0.703 0.842].

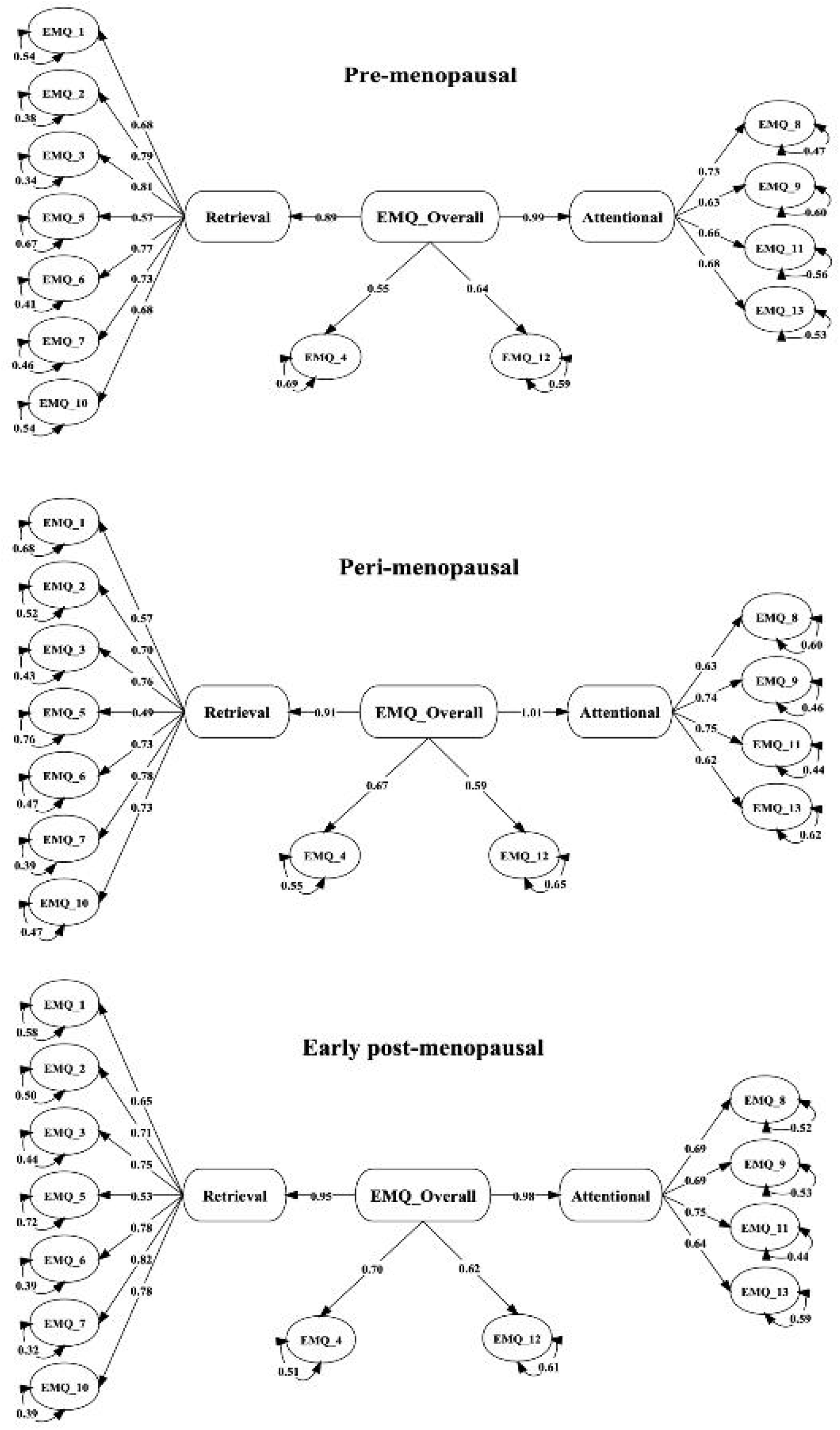

Based on the loaded items, as represented in Figure 2, the *Retrieval* factor was interpreted as the constellation of items concerning the failure of recalling recent events, retrieving words from memory and prospective memory, suggesting an information retrieval problem. The *Attentional* factor was defined as the set of items concerning track loss during conversations or readings revealing an attentional or working memory problem. The other two items (items 4 and 12) shared little in common and no clear interpretation of the processes involved in these items was available. Thus, these two items were not included for further analysis.

### Item-level analysis

Significant correlations were observed between retrieval subscale (correlation =0.96, *p* < 0.001), attentional subscale (r = 0.96, *p* < 0.001) and total scores (r = 0.75, *p* < 0.001). Furthermore, retrieval subscale and attentional subscale scores were significantly correlated with age (r = -0.13, *p* = 0.009 and r= -0.14, *p* = 0.003 respectively) and BMI (Body Mass Index) (r = 0.14, *p* = 0.003 and r = 0.14, *p* = 0.004 respectively).

From the one-way ANOVA and MANCOVA, three items (1,3,5) from the *Retrieval* factor as well as retrieval subscale score were found to be significantly different across groups. Other items (2,6,7,10) from *Retrieval* factor, all items (8,9,11,13) loading onto the *Attentional* factor, as well as attentional subscale score and total EMQ-R score were not significantly different across different menopausal groups. Detailed ANOVA and MANCOVA results were represented in Supplementary Table.

As shown in Figure 3, significant trends were identified in item 1 (‘Having to check whether you have done something you should have done’), item 3 (‘Forgetting that you were told something yesterday or a few days ago, and maybe having to be reminded about it’), and item 5 (‘Finding that a word is ‘on the tip of your tongue’. You know what it is but cannot quite find it’) between pre- and peri-menopausal groups, peri- and early post-menopausal groups, with a consistently highest mean score observed in the peri-menopausal group. Specifically, 46.9% of the women in the perimenopausal group reported that over the past months, item 5 has happened ‘once or more in a day’. This was followed by over one third (37.5%) of perimenopausal women reported experiencing item 1 ‘once or more in a day’. However, pre-menopausal women did not significantly differ with early postmenopausal women in these three items and a relatively similar mean score was present. Other items on *Retrieval* and *Attentional* all have a relatively lower prevalence (ranging from 18.7% to 1.8%) of daily complaints reported by women in all three groups.

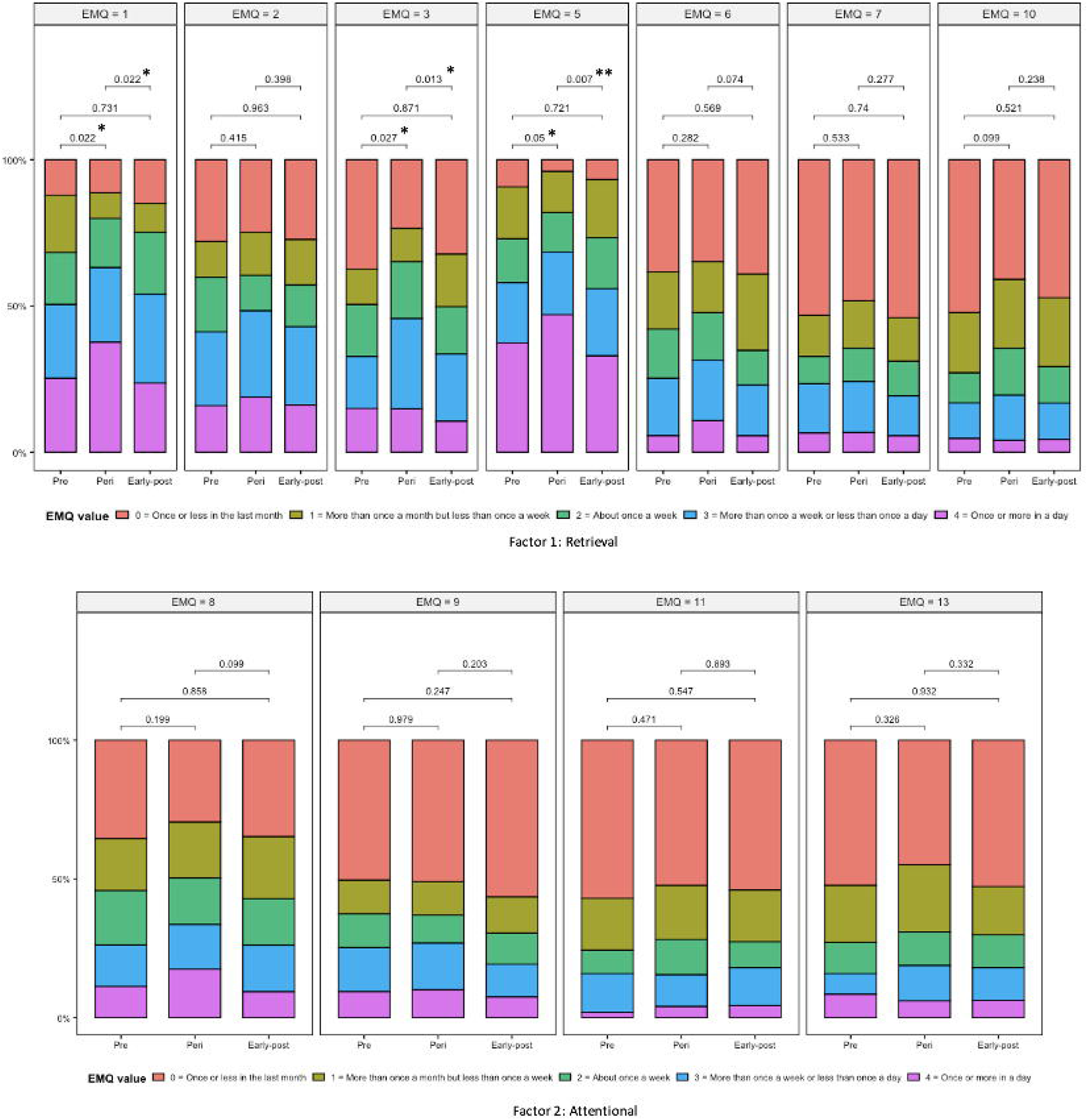

## DISCUSSION

The current study evaluated the factor structure of the EMQ-R in women across the menopause transition and investigated which cognitive complaints were commonly reported by women. The hypothesized CFA model has been validated through Cronbach test and indicated that the bifactor structure (*Retrieval* and *Attentional*) of the EMQ-R has good psychometric properties in pre-, peri- and early post-menopausal women. The peri-menopausal group reported increased retrieval difficulties, relative to both the pre-menopause and early post-menopause groups, suggesting a possible transition-related memory retrieval deficit during peri-menopause.

Consistent with the findings in multiple sclerosis population and healthy adults^23^, the bifactor model of the EMQ-R has a good fit and internal consistency in menopausal populations. The individual EMQ-R items and two factors (*Retrieval* and *Attentional*) identified have also indicated a good face validity for measuring brain fog. The EMQ-R could be potentially used as a standardized instrument to measure brain fog during menopause and future research may conduct a more comprehensive psychometric evaluation of the EMQ-R in menopausal population including convergent validity, known-groups validity, stability, or responsiveness.

### Memory retrieval complaints in menopause

The findings of the highest mean score in peri-menopausal group compared to pre- and early post-groups on three retrieval items and retrieval subscale are consistent with the increased memory complaints reported by perimenopausal women in previous research.^4,14^ A similar score between pre- and early post-suggests a temporary increase of memory retrieval difficulties during peri-menopausal stage, with a possible improvement to pre-menopausal level when entering into a post-menopausal stage. This finding has further reinforced the evidence of non-linear cognitive change trajectory across perimenopause as a result of transition stage effect and hormonal fluctuations.^13,33^

Items related to lexical or verbal information retrieval were the most commonly reported cognitive symptoms by peri-menopausal women. For instance, about half of peri-menopausal women have experienced ‘Tip of the tongue’ once or more in a day. Moreover, pre- and early post-menopausal groups did not significantly differ in these verbal retrieval related items, indicating that verbal memory, especially verbal retrieval/recall, may be particularly vulnerable to the menopause transition. This finding is consistent with the observed link between subjective memory and objective verbal recall from cross-sectional data^3,6^, as well as the decreased immediate and delayed verbal recall along the menopause transition identified in longitudinal study and associated with hormonal changes (e.g., dehydroepiandrosterone sulfate, DHEAS).^34^ Another longitudinal study examining verbal episodic memory (i.e., the ability to remember new information related to events or contextual information) reported an inverse association between follicle-stimulating hormone (FSH) and luteinizing hormone (LH) and memory performance in menopausal women.^35^

Other items in *Retrieval* did not yield a high frequency of daily and weekly reporting or any significant difference across groups. One reason is that all these items describe the phenomenon of forgetting to do something that targets more on prospective memory (e.g., completely forgetting to do things you planned to do) rather than verbal memory (neither verbal information retrieval nor verbal episodic memory), which further supports that menopausal-related cognitive change may be particularly related to specific type of verbal memory deficit. Future research may include the objective measures of prospective memory (e.g., event-based or time-based episodic memory task) to further study whether there are any changes in prospective memory during the menopause transition.

### Attentional complaints in menopause

The lack of reported complaints within the *Attentional* factor subscale in the peri-menopausal group are inconsistent with reduced attention/concentration performances either objectively^3,9,33^ or subjectively^3,36^ identified in previous studies. These items describe the encoding process that primarily relies on attentionally mediated cognitive processes requiring greater concentration effort (e.g., being unable to follow the thread of a story when reading). One possible reason for the inconsistency can be the different questions used in the current study and previous studies. In previous research, subjective attention was mostly measured by a single item question that directly asking the individual’s concern about general attention (e.g., Do you have problems with attention/concentration?), whereas the items involved in the EMQ-R used in the present study were more related to an exact daily phenomenon as described earlier. In this case, participants would be more likely to report that they had problems with attention as long as they had experience with any one of the attention-related problems. Although the attentional subscale has included four different items to capture daily attention problems, given that the item is describing a more specific behavior/circumstance, it is possible that these items could not fully cover or accurately depict the problems that each participant encounters in their daily lives. Thus, a principal component analysis on a longer version of the EMQ, covering more items describing daily attention problems (e.g., EMQ-R 20 or 28 items) would be a potential direction for future research to investigate the measurement of brain fog in menopausal population.

Another potential reason to account for the inconsistent results is that the items in the EMQ-R attentional subscale are targeting simple attentional process (i.e., concentrating when encoding information) instead of complex attentional process (i.e., controlling, shifting, or dividing attentional focus that allows for the manipulation when encoding information) involved in the objective measures used in previous studies^3,33^, such as Digit Span and California Computerized Assessment Package Reaction Time. This may suggest that the menopause-related attention change is more evident in complex attention rather than simple attention. Future research can further investigate the relationship between menopause and different types of attention.

### Limitations and future research

As discussed in previous sections, one potential limitation relating to the content of 13 item EMQ-R is that the attentional subscale may not capture more complex attention-related problems that perimenopausal women may experience in daily life. Moreover, some of the manifestations of menopausal brain fog defined recently by Maki and Jaff^11^, such as difficulty encoding words or numbers or switching between the tasks, are also not captured by the 13 item EMQ-R. Given that significant changes on verbal learning, working memory and psychomotor function during menopause transition have been reported with objective cognitive measures^33^, the EMQ-R may fall short of capturing all aspects of brain fog or cognitive complaints. Future research combining the EMQ-R with objective neuropsychological testing and possibly neuroimaging could help improve understanding of how these perceived/subjective deficits measured by the EMQ-R are associated with neuropsychological performance as well as better understand the aetiology of the retrieval problems identified in the current study.

Another key limitation is that the analysis and results interpretation of this study came from cross-sectional data, which can only allow for inferences about perceived deficits getting worse for an individual during peri-menopause and then improving during post-menopause. Therefore, a longitudinal study may help provide the evidence on subjective cognitive changes and brain fog characteristics across the menopausal transitional process.

### Potential clinical value

Given the significant cognitive complaints (i.e., brain fog) reported by women during menopausal transition, it is necessary to identify an easily implemented and validated self-reported scale to measure the level of subjective cognition difficulties in daily life. The bifactor structure of the Everyday Memory Questionnaire Revised (EMQ-R) has provided the scope to look at retrieval process and attentional process separately, which may provide better understanding of the underlying mechanism within the manifestation of brain fog and may lead to the development of more targeted treatment methods or coping strategies for women. Moreover, the items in the EMQ-R have provided detailed descriptions of daily memory and attention problem examples. By determining the prevalence and rating distribution of each item in the EMQ-R, women and clinicians would grasp a more direct overview of the most commonly reported symptoms during menopause in daily life, which would strengthen their understanding and knowledge in menopausal-related cognitive changes.

Importantly, the good psychometric properties (i.e., reliability) of the EMQ-R in menopausal populations has provided the initial evidence of the potential clinically diagnostic value of mental health in menopausal women and yielded valuable information to support the development of a cut off score of the EMQ-R to clinically differentiate those whose daily life functioning is greatly influenced by brain fog. A more standardized clinical diagnosis of brain fog can help women and clinicians to identify the potential risk of neurodegenerative diseases or dementia later in life at a very early stage. Currently, there is no available cutoff score for the EMQ-R used in menopausal population. Future research may consider the levels of menopausal-related complaints (e.g., quality of life, general well-being, vasomotor symptoms, psychosocial impacts, etc.) to further define the corresponding the EMQ-R cutoff score for the purpose of testing the criterion validity or predictive validity of the EMQ-R.

## CONCLUSIONS

In the current study, the EMQ-R has indicated good psychometric properties (i.e., fit and internal consistency) in a menopausal sample. Pre-, peri- and early post-menopausal groups did not significantly differ in overall subjective cognition (i.e., brain fog) and subjective attention. Significantly higher levels of memory retrieval complaints were observed in peri-menopausal women, as compared to pre- and post-menopausal women, with frequent reporting of verbal-related retrieval problems. This suggests that the trajectory of subjective cognitive changes across menopause may be non-linear and the decrease in memory retrieval may be temporary during peri-menopause with subjective cognition returning to pre-menopausal levels in post-menopausal years. This study has provided the preliminary evidence for the EMQ-R as a standardized instrument to measure brain fog during menopause which can further assist women and clinicians to understand cognitive change during menopause transition.

## Data Availability

All data produced in the present study are available upon reasonable request to the authors.

## Acknowledgments

The authors would like to thank the people who took time to comment on this review.

