## Supplemental Tables for "Evaluation of The Everyday Memory Questionnaire-Revised in Menopausal Population: Understanding the Brain Fog During Menopause"

| Supplementary Table. Descriptive Information for EMQ_R outcomes | | | | | | |
| --- | --- | --- | --- | --- | --- | --- |
| Item | Pre | Peri | Early-post | *F Statistics* | p-value | Test |
|  | *Mean (SD)* | *Mean (SD)* | *Mean (SD)* |  |  |  |
| *Factor 1: Retrieval* | *11.8 (7.36)* | *13.58 (6.99)* | *11.73 (7.17)* | *3.17* | ***0.043**** | *MANCOVA* |
| 1. Having to check whether you have done something that you should have done. | 2.32 (1.36) | 2.69 (1.36) | 2.38 (1.35) | 3.03 | **0.049*** | *ANOVA* |
| 2. Forgetting when it was that something happened; for example, whether it was yesterday or last week. | 1.89 (1.46) | 2.03 (1.48) | 1.89 (1.47) | 0.43 | 0.654 | *ANOVA* |
| 3. Forgetting that you were told something yesterday or a few days ago, and maybe having to be reminded about it. | 1.61 (1.50) | 2.02 (1.40) | 1.61 (1.41) | 3.89 | **0.021*** | *ANOVA* |
| 5. Finding that a word is 'on the tip of your tongue'. You know what it is but cannot quite find it. | 2.59 (1.39) | 2.93 (1.24) | 2.55 (1.31) | 3.77 | **0.024*** | *ANOVA* |
| 6. Completely forgetting to do things you said you would do, and things you planned to do. | 1.35 (1.32) | 1.55 (1.42) | 1.24 (1.29) | 2.08 | 0.127 | *ANOVA* |
| 7. Forgetting important details of what you did or what happened to you the day before. | 1.09 (1.38) | 1.18 (1.37) | 1.02 (1.31) | 0.56 | 0.571 | *ANOVA* |
| 10. Forgetting to tell somebody something important, perhaps forgetting to pass on a message or remind someone of something. | 0.96 (1.24) | 1.18 (1.24) | 1.03 (1.22) | 1.09 | 0.336 | *ANOVA* |
| *Factor 2: Attentional* | *4.53 (4.10)* | *5.01 (4.20)* | *4.65 (4.12)* | *1.05* | *0.350* | *MANCOVA* |
| 8. When talking to someone, forgetting what you have just said. Maybe saying 'what was I talking about?' | 1.48 (1.40) | 1.72 (1.48) | 1.43 (1.36) | 1.738 | 0.177 | *ANOVA* |
| 9. When reading a newspaper or magazine, being unable to follow the thread of a story; losing track of what it is about. | 1.21 (1.44) | 1.23 (1.47) | 1.01 (1.35) | 1.16 | 0.316 | *ANOVA* |
| 11. Getting the details of what someone was told you mixed up and confused. | 0.85 (1.17) | 0.95 (1.22) | 0.96 (1.26) | 0.29 | 0.748 | *ANOVA* |
| 13. Repeating to someone what you have just told them or asking some the same question twice. | 0.99 (1.31) | 1.11 (1.27) | 1.01 (1.30) | 0.32 | 0.728 | *ANOVA* |
| *Miscellaneous:* |  |  |  |  |  |  |
| 4. Starting to read something (a book or an article in a newspaper, or a magazine) without realizing you have already read it before. | 0.69 (1.14) | 0.78 (1.20) | 0.67 (1.18) | 0.35 | 0.704 | *ANOVA* |
| 12. Forgetting where things are normally kept or looking for them in the wrong place. | 0.61 (1.09) | 0.82 (1.23) | 0.74 (1.22) | 0.98 | 0.376 | *ANOVA* |
| *Total* | *17.64 (12.09)* | *20.19 (12.01)* | *15.55 (12.18)* | *2.22* | *0.110* | *ANOVA* |
| SD = Standard Deviation.  EMQ_R, Everyday Memory Questionnaire_Revised. * indicated p <0.05. | | | | | | |
